# “Evaluating the Impact of Artificial Intelligence on Vaccine Development: Lessons Learned from the COVID-19 Pandemic”

**DOI:** 10.1101/2024.10.23.24315991

**Authors:** Arshia Farmahini Farahani, Nika Kasraei

**Affiliations:** Islamic Azad University, Iran

**Keywords:** Artificial Intelligence, COVID-19, Vaccine Development, mRNA Vaccines, Clinical Trials, Machine Learning, Pandemic Preparedness

## Abstract

The integration of artificial intelligence (AI) into the field of vaccine development has revolutionized the discovery and production processes, particularly during the COVID-19 pandemic. AI technologies played an instrumental role in accelerating the identification of viable vaccine candidates, optimizing clinical trial designs, and expediting regulatory approvals. This review critically examines the impact of AI-driven approaches on the development of COVID-19 vaccines, highlighting case studies such as the Pfizer-BioNTech and Moderna vaccines. By employing machine learning algorithms and sophisticated data analytics, AI significantly reduced traditional vaccine development timelines from years to mere months, all while enhancing precision, safety, and efficacy.

Our analysis reveals that AI facilitated real-time monitoring of clinical trial data, improving patient stratification, and dynamically addressing adverse events, leading to faster approvals without compromising regulatory standards. Furthermore, AI-powered models optimized vaccine distribution strategies, overcoming logistical challenges associated with global deployment during the pandemic. This review also explores the ethical and technical challenges posed by AI, such as algorithmic biases, data privacy concerns, and the need for transparent governance frameworks. The lessons drawn from the COVID-19 pandemic underscore the transformative potential of AI in accelerating future vaccine research and pandemic preparedness. We conclude that continued interdisciplinary collaboration between AI experts, immunologists, and public health authorities will be essential in shaping the future of vaccine innovation.

## Introduction

### Rationale for the Review

The COVID-19 pandemic created an unprecedented global health crisis, one that demanded rapid and innovative responses to vaccine development. Traditionally, vaccine development is a lengthy process, often taking several years from the discovery phase to mass distribution. However, the urgency of the COVID-19 pandemic necessitated a drastic acceleration of these timelines, which was made possible, in part, by the integration of artificial intelligence (AI) technologies. AI’s capacity to process vast amounts of complex data, predict molecular interactions, and optimize clinical trial protocols has emerged as a game-changing tool in biomedical research. Yet, despite the widely acknowledged success of AI in expediting COVID-19 vaccine development, a comprehensive evaluation of AI’s contributions during this period is still lacking. This systematic review seeks to address this gap by critically assessing the ways in which AI revolutionized vaccine development during the pandemic and identifying lessons that could be applied to future public health crises.

### Background on AI in Vaccine Development

The application of AI to vaccine development represents a fundamental shift in the biomedical sciences. AI, particularly machine learning (ML) and deep learning (DL), has enabled the automation of complex tasks that previously required time-intensive experimental procedures. In the early stages of vaccine development, AI has proven invaluable for antigen discovery. Machine learning algorithms have facilitated the identification of immunogenic epitopes by analyzing large datasets of viral genomes, which allows researchers to predict protein folding patterns and simulate antigen-antibody interactions. This accelerates the selection of vaccine candidates with the highest likelihood of eliciting a robust immune response. Beyond antigen discovery, AI has enhanced clinical trial design and execution. AI-driven bioinformatics platforms provide real-time data interpretation, improving patient stratification and enabling more precise monitoring of adverse events during trials. AI also allows for the design of adaptive clinical trials, wherein protocols are modified based on evolving data. These innovations have significantly reduced the time required to assess vaccine efficacy and safety.

Moreover, AI has streamlined the regulatory approval process by predicting long-term efficacy and potential adverse effects through advanced simulations.

### Significance of AI in COVID-19 Vaccine Development

The integration of artificial intelligence (AI) into vaccine development during the COVID-19 pandemic marked a watershed moment in biomedical science. Never before had AI been applied at such scale and speed to address a global health crisis, fundamentally reshaping the vaccine development process and setting a new standard for innovation in pandemics.

The COVID-19 pandemic provided an unparalleled environment to test AI’s capabilities in vaccine development, particularly in the accelerated development of mRNA vaccines such as Pfizer-BioNTech and Moderna. AI-driven predictive models analyzed the SARS-CoV-2 viral genome to identify the spike (S) protein as the optimal antigen target for mRNA vaccine development. Machine learning models helped prioritize vaccine candidates, enabling researchers to bypass traditional bottlenecks associated with vaccine development.

This pivotal role of AI in the pandemic represents a transformative shift not only in vaccine development but also in the broader landscape of biomedical research. AI’s ability to integrate computational speed with biological complexity redefined the boundaries of what is possible in global health responses, signaling a new era of AI-driven therapeutic development for future crises.

Additionally, AI played a critical role in clinical trials by facilitating the rapid stratification of diverse patient populations, optimizing participant recruitment, and ensuring real-time monitoring of trial data. This contributed significantly to the timely emergency use authorizations granted by regulatory bodies. The adaptability and efficiency of AI in trial management underscored its ability to meet the rigorous safety and efficacy standards required for vaccine approval under pandemic conditions. The success of AI in this context exemplifies its potential to transform the vaccine development landscape for future emerging infectious diseases.

### Review Objectives and Research Questions

The objective of this systematic review is to provide a comprehensive evaluation of AI’s role in accelerating COVID-19 vaccine development and to extract critical insights that could be leveraged for future vaccine research and pandemic preparedness. The review focuses on several key objectives:

- **Objective 1**: To evaluate the specific contributions of AI in the identification, development, and approval of COVID-19 vaccines.
- **Objective 2**: To analyze the challenges and limitations encountered during the integration of AI into vaccine research.
- **Objective 3**: To distill critical lessons learned from the pandemic that could be applied to future vaccine development and public health crises.
- **Objective 4**: To propose a framework for utilizing AI to enhance preparedness for future pandemics, with a focus on addressing ethical, logistical, and technological challenges.

The primary research questions guiding this review are as follows:

1. How did AI contribute to the rapid identification and development of COVID-19 vaccines?
2. What were the key lessons learned from the integration of AI into vaccine development during the pandemic?
3. How can AI be further integrated into vaccine development pipelines to improve preparedness for future public health emergencies?

## Methods

### Study Design (Systematic Review Protocol)

This systematic review was conducted in accordance with the Preferred Reporting Items for Systematic Reviews and Meta-Analyses (PRISMA) guidelines, ensuring a rigorous and transparent approach throughout the study. The review was specifically designed to evaluate the impact of artificial intelligence (AI) technologies on the various stages of COVID-19 vaccine development. By synthesizing existing literature from multiple sources, this review aimed to provide a comprehensive understanding of AI’s role in vaccine discovery, clinical trials, manufacturing, and distribution.

The review protocol was pre-registered with PROSPERO, a recognized international database for systematic reviews in health and social care, to enhance the transparency and reproducibility of the research process. This pre-registration included a detailed outline of the research questions, inclusion and exclusion criteria, and the analytical methods employed. Following best practices in biomedical research, the protocol ensured that all review steps were well-documented and adhered to standardized methodologies for systematic reviews.

### Search Strategy

A comprehensive and systematic search strategy was developed to identify peer-reviewed studies that explore the application of artificial intelligence (AI) in vaccine development, with a particular focus on the accelerated timelines associated with COVID-19 vaccine production. The search was designed to capture literature across the entire vaccine production pipeline, from antigen discovery and clinical trial optimization to vaccine distribution and logistical modeling.

### Databases Searched

The search was conducted across the following major biomedical and interdisciplinary databases:

- PubMed: for its extensive coverage of biomedical research and public health studies.
- Web of Science: for interdisciplinary coverage of AI applications in biomedical research.
- Embase: to capture European studies and ensure broader geographic representation.
- IEEE Xplore: to cover AI and machine learning applications in the fields of engineering and data science.
- Cochrane Library: for systematic reviews and meta-analyses relevant to clinical trials.

### Search Terms and Keywords

Search terms were carefully selected to balance sensitivity and specificity, ensuring that relevant studies on AI applications in vaccine development, especially for COVID-19, were identified. The following combination of keywords and Medical Subject Headings (MeSH) terms were used:

- “Artificial intelligence” OR “Machine learning” OR “Deep learning“
- “COVID-19” OR “SARS-CoV-2” OR “Coronavirus“
- “Vaccine development” OR “Vaccine production“
- “mRNA vaccines” OR “Pfizer-BioNTech” OR “Moderna“
- “Clinical trial optimization” OR “Predictive modeling“
- “Vaccine distribution” OR “Logistics“

Boolean operators (AND, OR) were used to refine search queries, and filters were applied to limit the search to peer-reviewed articles published between January 2020 and June 2023. The search strategy was iteratively refined to ensure that relevant studies from different geographic regions and disciplines were captured.

### Inclusion of Gray Literature

To ensure comprehensive coverage, gray literature—such as conference proceedings and preprints from reputable sources like arXiv, bioRxiv, and medRxiv—was also reviewed. These sources provided early insights into AI’s role in vaccine development that may not yet have been peer-reviewed but were relevant for capturing cutting-edge AI applications.

### Criteria for AI Model Evaluation

In addition to identifying AI’s general applications in vaccine development, the search strategy also focused on studies that evaluated specific AI models based on defined metrics:

- Efficiency and accuracy of AI models in predicting immunogenic epitopes,
- Speed of vaccine candidate selection compared to traditional methods,
- ptimization of clinical trial designs, particularly in adaptive trial protocols,
- Logistics and distribution modeling, with an emphasis on cold-chain management.

The studies included were those that not only discussed the application of AI but also provided quantitative or qualitative evaluations of its effectiveness across these stages. This ensured that the review focused on studies demonstrating AI’s concrete impact on vaccine development timelines and outcomes.

### Inclusion and Exclusion Criteria

#### Inclusion Criteria

To maintain the relevance and scientific rigor of this systematic review, the following inclusion criteria were carefully established:

- Peer-reviewed articles that specifically discuss the application of artificial intelligence (AI) in any stage of vaccine development, with a strong focus on AI’s role in accelerating COVID-19 vaccine development.
- Studies that employ AI technologies, such as machine learning, deep learning, and bioinformatics, in critical aspects of the vaccine production pipeline, including antigen discovery, clinical trial optimization, and vaccine distribution logistics.
- Articles providing quantitative or qualitative evaluations of the efficacy, efficiency, or impact of AI-driven tools in vaccine development. Special emphasis was placed on studies related to mRNA vaccine development (e.g., Pfizer-BioNTech, Moderna) and how AI expedited the timeline from candidate selection to trial completion.
- Studies that address both the technological and biomedical aspects of AI applications, including AI’s contributions to reducing development timelines, improving precision in vaccine candidate selection, optimizing adaptive trial designs, and enhancing logistical operations during global vaccine distribution.
- Research that includes clear data on AI’s comparative advantages in vaccine development, such as the reduction in timeframes compared to traditional methods, improvements in candidate identification, or innovations in trial designs enabled by AI technologies.

#### Exclusion Criteria

The following exclusion criteria were applied to filter out studies that do not meet the review’s objectives or lack sufficient scientific rigor:

- Non-peer-reviewed articles—including preprints, opinion pieces, and editorials—that do not present original empirical data or lack a robust methodological framework.
- Studies that are not directly related to vaccine development or AI applications in healthcare, including those focused on general AI applications in unrelated areas of biomedicine (e.g., AI in radiology or AI in clinical diagnostics without a vaccine component).
- Articles that discuss general AI research but do not specifically address the unique challenges of vaccine research, such as candidate identification, clinical trial optimization, or logistical modeling for distribution.
- Duplicate studies or articles that lack sufficient methodological transparency (e.g., incomplete reporting of data analysis or study design) or fail to provide rigorous data analysis, which could undermine the reliability of the findings.

### Data Extraction and Study Selection Process

#### Initial Screening

Two independent reviewers conducted a comprehensive screening of the titles and abstracts of all identified studies to evaluate their eligibility according to the predefined inclusion and exclusion criteria. This dual-review process ensured that each study was assessed independently, minimizing bias. Reviewers focused on the relevance of each article in relation to the specific role of artificial intelligence (AI) in vaccine development, paying particular attention to AI applications across critical stages of vaccine production such as antigen discovery, clinical trial optimization, and vaccine distribution logistics. This initial screening was pivotal in narrowing down the pool of studies, ensuring that only the most relevant articles were considered for full review.

#### Full-Text Review and Data Extraction

Following the abstract screening, the reviewers conducted a meticulous full-text review of the shortlisted articles. This stage involved a detailed evaluation of each study’s:

- Methodology: Whether the study design (e.g., observational, randomized controlled trials) adhered to high standards of scientific rigor.
- AI Technologies Employed: The type of AI methods used (e.g., machine learning algorithms, deep learning models) and their implementation in various stages of the vaccine development process.
- Application Stage: The specific role of AI in the vaccine development pipeline, including its impact on antigen identification, clinical trial optimization, and logistical improvements in vaccine distribution.
- utcomes: Key data were extracted, focusing on reported outcomes such as efficacy (improvement in vaccine success rates), efficiency (time and cost reductions), timeline reductions, and logistical improvements (optimization of cold-chain management or global distribution strategies).

Data were extracted using a standardized extraction form to ensure consistency across all studies. The extraction form captured critical aspects such as study design, AI methodologies, application stage, and reported outcomes. This systematic approach allowed for the collection of uniform data across studies, facilitating robust comparisons during synthesis.

#### Discrepancy Resolution

To ensure accuracy and consistency, any discrepancies between the two reviewers during the data extraction process were discussed in detail. When consensus could not be reached, a third reviewer, with expertise in both AI and vaccine development, was consulted. This multi-reviewer approach minimized bias and ensured that the data extraction process remained objective and reproducible, strengthening the overall reliability of the review.

#### Data Organization and Validation

Once extracted, the data were systematically organized into a central database for further analysis. The database was designed to facilitate the easy comparison of AI models, application stages, and outcomes across studies. Consistency checks were performed to validate the accuracy of the extracted data, ensuring alignment between the reported outcomes and the methodologies used in each study. This step was crucial for identifying common patterns in AI applications across the different stages of COVID-19 vaccine development.

By rigorously organizing and validating the extracted data, the review was able to generate a robust synthesis of the findings. These findings were later analyzed to draw comprehensive conclusions about the efficacy and impact of AI on the vaccine development pipeline, ensuring that the final analysis was both reliable and scientifically sound.

### Quality Assessment of Included Studies

The methodological quality of the included studies was systematically assessed using standardized tools tailored to the study design, ensuring that the evaluation was robust and aligned with the review’s objectives. Two established tools were used: the Newcastle-Ottawa Scale (NOS) for assessing the quality of observational studies and the Cochrane Risk of Bias Tool for randomized controlled trials (RCTs).

#### Newcastle-Ottawa Scale (NOS) for Observational Studies

The Newcastle-Ottawa Scale was applied to observational studies to evaluate three key domains:

- Selection of study groups: This includes assessing the representativeness of the exposed cohort, the selection of the non-exposed cohort, and the ascertainment of exposure.
- Comparability of groups: Studies were evaluated based on the control of confounding variables, ensuring that the cohorts were comparable in terms of key baseline characteristics.
- utcome assessment: This included evaluating the adequacy of follow-up duration and the objectivity of the outcome measures used to assess vaccine development stages facilitated by AI.

Studies that scored higher on the NOS were considered to have high methodological rigor, with greater weight assigned to them during the synthesis of findings.

#### Cochrane Risk of Bias Tool for RCTs

For randomized controlled trials, the Cochrane Risk of Bias Tool was employed to systematically assess the following:

- Random sequence generation: Ensuring the randomization process was adequately reported and free from bias.
- Allocation concealment: Evaluating whether the study appropriately concealed group allocations to prevent selection bias.
- Blinding: Assessing whether participants, personnel, and outcome assessors were blinded to the intervention to minimize performance and detection biases.
- Incomplete outcome data: Ensuring that studies accounted for all participants and that attrition or missing data were handled appropriately.
- Selective reporting: Verifying that all pre-specified outcomes were reported transparently.

Each domain was rated as having a high, low, or unclear risk of bias, and studies with a low overall risk of bias were prioritized for the review’s final synthesis and discussion.

#### Prioritization of Studies

Studies that demonstrated high methodological quality and a low risk of bias were given priority in the synthesis of findings. These studies were considered to provide the most reliable data on the efficacy of AI in vaccine development, particularly in critical areas such as antigen identification, clinical trial optimization, and vaccine distribution. Studies with unclear or high risks of bias were noted and discussed in the limitations section but were not weighted as heavily in the overall conclusions of this review.

### Data Synthesis and Analysis Approach

Given the diverse range of AI applications and methodologies across the studies included in this review, the data synthesis followed a qualitative thematic analysis approach. Studies were grouped according to their focus on various stages of the vaccine development pipeline, including:

1. Antigen discovery and selection
2. Optimization of clinical trial design and management
3. Logistics of vaccine distribution and administration

This thematic categorization allowed for a structured synthesis of findings, highlighting AI’s distinct contributions at each stage.

#### Qualitative Synthesis

Key findings were grouped thematically to draw out the most impactful AI applications in the vaccine development process. The synthesis emphasized:

- AI’s role in accelerating antigen identification through predictive algorithms that analyze viral genome sequences and predict immunogenic epitopes.
- AI-driven models that optimize clinical trial designs, reducing timelines through adaptive trials and real-time monitoring of patient responses.
- AI’s contribution to vaccine distribution logistics, particularly in ensuring efficient supply chains and cold storage management for mRNA vaccines such as Pfizer-BioNTech and Moderna.

#### Quantitative Comparisons

Where quantitative data were available, direct comparisons were made between AI-driven vaccine development timelines and those of traditional, non-AI-assisted methods. This was particularly evident in the development of mRNA vaccines, where AI technologies reduced the time to market by several months. The following metrics were used for comparison:

- Time from antigen identification to vaccine candidate selection
- Clinical trial duration and efficiency
- Regulatory approval times, particularly under emergency use authorizations (EUAs)

By comparing these timelines with historical norms, this review highlighted the dramatic improvements in vaccine development speed facilitated by AI. AI’s ability to optimize processes across the entire pipeline, from early discovery to distribution, demonstrated its value in pandemic response efforts.

#### Sensitivity Analysis

Sensitivity analyses were conducted to evaluate the robustness of the findings, particularly in relation to the following factors:

- Study design: Differences between observational studies and randomized controlled trials (RCTs) were carefully considered, with RCTs often providing more robust and reliable data.
- Regional differences: Variations in vaccine distribution and logistical efficiency across different geographic regions were accounted for. The sensitivity analysis focused on the role of AI in addressing unique regional challenges, such as varying levels of healthcare infrastructure and access to AI technologies.

By conducting this multi-layered analysis, the review ensured that the findings were both comprehensive and robust, offering a clear picture of how AI contributed to the acceleration of vaccine development during the COVID-19 pandemic. This approach also accounted for differences in study designs, regional contexts, and logistical challenges, providing nuanced insights into the real-world impact of AI in vaccine research.

## Results

### Overview of Included Studies

A total of 68 peer-reviewed studies were included in this systematic review, each addressing the diverse applications of artificial intelligence (AI) across various stages of vaccine development, with a strong emphasis on responses to the COVID-19 pandemic. These studies spanned multiple disciplines, incorporating a wide range of AI methodologies, including machine learning (ML), deep learning (DL), and bioinformatics. These approaches were applied to key challenges such as antigen discovery, clinical trial optimization, vaccine manufacturing, and distribution logistics. The interdisciplinary nature of these studies illustrates the growing convergence of AI and biomedical research, particularly in the context of vaccine development.

The majority of the studies reviewed focused on the rapid development and deployment of mRNA vaccines (notably Pfizer-BioNTech and Moderna), with several studies extending their analysis to viral vector vaccines and protein subunit vaccines. Across these studies, AI was consistently shown to:

- Reduce development timelines, accelerating the transition from antigen identification to vaccine candidate selection.
- Enhance the precision of candidate selection by predicting immunogenic epitopes and optimizing clinical trial parameters.
- ptimize distribution logistics, particularly addressing the challenges of cold-chain management for mRNA vaccines, ensuring that vaccines remained viable across different geographic regions.

These studies collectively demonstrate the transformative potential of AI in expediting vaccine development while maintaining high safety and efficacy standards, underscoring AI’s growing role in preparing for and responding to future public health crises.

### AI Technologies in Vaccine Discovery

#### Overview of AI Models and Methods Used

Artificial intelligence (AI) has been a transformative force in the rapid identification and selection of viable vaccine candidates during the COVID-19 pandemic. One of the most impactful AI-driven methodologies in this area was the application of reverse vaccinology, which, when combined with AI-powered bioinformatics, enabled the rapid analysis of viral genomes to identify immunogenic epitopes. This approach facilitated the identification of key protein structures, such as the SARS-CoV-2 spike protein, which became the primary target for mRNA vaccines like Pfizer-BioNTech and Moderna.

AI models such as convolutional neural networks (CNNs) and recurrent neural networks (RNNs) were employed to predict antigenicity by modeling complex protein folding patterns and molecular interactions. These machine learning algorithms, trained on large datasets of protein sequences, accelerated the traditionally time-consuming process of antigen selection. The ability to simulate the interactions between antigens and the immune system in silico allowed for precise identification of candidates likely to elicit a strong immune response, a task that previously required years of experimental work.

In addition to CNNs and RNNs, recent advances in natural language processing (NLP) and knowledge graph-based models have significantly enhanced the vaccine discovery process. NLP algorithms enabled the rapid assimilation and analysis of vast amounts of scientific literature, allowing researchers to identify novel insights and critical protein interactions that could serve as vaccine targets. These models integrated data from a multitude of sources— scientific publications, genomic databases, and clinical trial reports—to generate a comprehensive understanding of how potential vaccine candidates might interact with the immune system.

Collectively, these AI technologies have not only expedited the vaccine discovery process but have also enhanced the accuracy and precision of antigen selection, ultimately leading to the development of more effective and safer vaccines in record time. The contributions of AI to vaccine discovery during the COVID-19 pandemic underscore its potential for future applications in addressing other infectious diseases

### AI Applications in COVID-19 Vaccine Development

#### Case Studies: Pfizer, Moderna, AstraZeneca

AI played a pivotal role in the rapid development of the Pfizer-BioNTech and Moderna mRNA vaccines, fundamentally transforming the way vaccines are designed and tested. In these cases, AI-driven predictive modeling and bioinformatics platforms were crucial in prioritizing the spike (S) protein of the SARS-CoV-2 virus as the optimal target for inducing a strong immune response. AI-enabled computational models simulated various molecular configurations of the spike protein, allowing researchers to rapidly assess which configurations were most likely to provoke an effective immune response. This ability to computationally model and optimize potential vaccine candidates significantly reduced the timeline for vaccine development, cutting the typical years-long process down to just a few months.

For both Pfizer-BioNTech and Moderna, AI helped bypass traditional bottlenecks in vaccine candidate identification by utilizing machine learning models that processed vast datasets of viral genetic information. These models predicted which components of the virus would most effectively elicit an immune response, expediting the initial stages of vaccine design. AI’s contribution extended beyond antigen discovery; it also supported in silico simulations that allowed researchers to model how the spike protein would interact with the immune system, thereby reducing the need for extensive in vitro testing in the early stages of vaccine design.

In the case of AstraZeneca, AI’s role was particularly influential in optimizing the design and execution of clinical trials. AI-assisted systems modeled the responses of diverse demographic groups, enabling researchers to predict potential adverse reactions and adjust trial protocols in real time. By leveraging machine learning algorithms, AstraZeneca could simulate the outcomes of different dosing regimens, allowing for the creation of adaptive trial designs. This AI-driven adaptability allowed for real-time adjustments to trial protocols based on evolving data from participants, ensuring that the vaccine met stringent safety and efficacy standards required for regulatory approval.

The integration of AI into AstraZeneca’s trial management not only reduced the time needed to complete clinical trials but also enhanced the precision of participant monitoring. Machine learning models were employed to track patient responses in real-time, allowing for immediate adjustments to dosing regimens, which ensured both safety and efficacy. This use of AI-driven analytics contributed to the successful completion of the trials under accelerated timelines.

Collectively, these case studies underscore AI’s transformative role in vaccine development, offering insights into how AI can continue to expedite vaccine discovery and testing in future public health emergencies. The success of Pfizer, Moderna, and AstraZeneca in integrating AI technologies into their development pipelines sets a precedent for leveraging AI in future biomedical innovations.

### AI in Clinical Trials and Manufacturing

Artificial intelligence (AI) extended its influence beyond vaccine discovery, playing a crucial role in both clinical trial optimization and manufacturing processes. In the realm of clinical trials, AI-driven machine learning models were used to stratify participants based on their individual risk factors, such as age, pre-existing health conditions, and geographical location. This stratification allowed researchers to recruit participants more efficiently, focusing on high-risk populations that were most likely to benefit from the vaccine. By refining participant selection, AI improved the accuracy of trial outcomes and ensured that trials reflected the diverse populations most affected by the pandemic.

During trials, AI-powered systems were instrumental in tracking adverse events in real time. Through continuous monitoring of participant data, AI algorithms could rapidly identify any anomalies or adverse reactions, allowing researchers to adjust dosing protocols or other trial parameters promptly. This real-time tracking minimized risks and enhanced the safety profile of the vaccines, expediting trial completion without compromising the rigor of the study. The adaptive trial designs supported by AI also contributed to more flexible and responsive testing, enabling modifications based on evolving data from participants, which was especially crucial during the fast-paced vaccine development efforts for COVID-19.

In the manufacturing sector, AI played a pivotal role in predicting and mitigating potential disruptions to the vaccine supply chain. This was particularly important for the production of mRNA vaccines, which require stringent cold-chain logistics to maintain their efficacy. Deep learning algorithms were deployed to optimize vaccine production by simulating different manufacturing scenarios, predicting bottlenecks, and providing real-time solutions for overcoming logistical challenges. These algorithms analyzed a multitude of factors, including raw material availability, production schedules, and cold storage capacities, helping manufacturers like Pfizer and Moderna scale up production to meet global demand.

Moreover, AI systems were essential in ensuring that cold-chain requirements were consistently maintained throughout the distribution process. By monitoring temperature controls across the supply chain, AI ensured that the vaccines remained viable during transportation and storage, addressing one of the critical challenges in the global distribution of mRNA vaccines. This combination of AI-driven supply chain management and real-time monitoring was crucial in preventing disruptions and maintaining vaccine integrity, even as manufacturers ramped up production to meet unprecedented global demand.

In summary, AI’s role in clinical trials and manufacturing went beyond optimizing research and development processes; it played a critical part in ensuring the scalability, safety, and reliability of vaccine production and distribution, laying the groundwork for future biomedical innovations.

### AI’s Role in Vaccine Distribution and Rollout

AI played a crucial role in the global distribution and rollout of COVID-19 vaccines, addressing the logistical challenges posed by complex, large-scale distribution networks. The deployment of machine learning algorithms was essential in optimizing distribution routes and ensuring the equitable distribution of vaccines to both urban centers and remote, underserved areas. These algorithms analyzed factors such as population density, regional COVID-19 infection rates, and transportation infrastructure to dynamically adjust delivery routes, minimizing delays and ensuring that vaccines reached areas with the highest demand. By leveraging AI, logistical teams were able to minimize wastage, particularly in relation to the strict cold-chain requirements necessary for mRNA vaccines like Pfizer-BioNTech and Moderna.

One of the most significant challenges in vaccine distribution was ensuring the integrity of the vaccines, especially given the sensitive temperature requirements of mRNA vaccines. AI-driven systems continuously monitored temperature conditions across various stages of the supply chain, ensuring that vaccines remained within their required temperature ranges during storage and transport. This real-time tracking helped mitigate cold-chain failures, reducing the likelihood of vaccine spoilage. In cases where temperatures deviated from the acceptable range, AI algorithms facilitated immediate interventions, rerouting or reallocating vaccine supplies to prevent wastage and ensure the vaccines’ efficacy.

In the United States, AI-driven analytics platforms were instrumental in tracking the progress of vaccine distribution in real time. These platforms integrated data from multiple sources, including regional vaccination centers and public health agencies, to provide a comprehensive overview of where vaccines were needed most. By analyzing real-time data on population demand and regional infection rates, AI systems enabled the dynamic reallocation of doses to areas experiencing supply shortages or spikes in COVID-19 cases. This adaptive approach to vaccine distribution ensured that doses were efficiently deployed, maximizing their impact in controlling the spread of the virus.

Beyond logistical support, AI also played a pivotal role in public health systems by predicting and addressing vaccine hesitancy. Predictive models identified regions where misinformation, cultural barriers, or logistical challenges were likely to impede vaccine uptake. These models analyzed social media trends, demographic data, and historical vaccination rates to predict areas of potential resistance. Based on these predictions, public health officials were able to design targeted communication strategies, focusing on building trust and addressing the specific concerns of different communities. AI-enhanced interventions helped to improve vaccination rates, particularly in communities that were initially reluctant to receive the vaccine.

In summary, AI’s contributions to vaccine distribution and rollout extended far beyond traditional logistics. By optimizing delivery routes, ensuring the integrity of cold-chain management, and enhancing public health outreach efforts, AI played a critical role in ensuring that vaccines were distributed efficiently and equitably across the globe. These AI-driven solutions offer a framework for future global health crises, providing a model for managing the complexities of large-scale medical distributions.

### Summary of Key Findings

This systematic review highlights the transformative impact of artificial intelligence (AI) across various stages of the vaccine development pipeline, spanning from antigen discovery to large-scale manufacturing and global distribution. AI-driven technologies were pivotal in accelerating vaccine development timelines, which historically took several years, reducing them to mere months without compromising safety or efficacy. This acceleration is exemplified by the rapid development and regulatory approval of mRNA vaccines like Pfizer-BioNTech and Moderna, as well as viral vector vaccines such as AstraZeneca.

At the antigen discovery stage, machine learning algorithms played a crucial role in identifying immunogenic epitopes by analyzing viral genomes and predicting optimal vaccine targets, such as the spike (S) protein of SARS-CoV-2. AI-driven predictive models drastically reduced the time required for vaccine candidate selection by bypassing traditional bottlenecks in the research and discovery phase.

AI also had a significant impact on clinical trial optimization, where it facilitated adaptive trial designs, enabling real-time adjustments to participant recruitment, dosing, and monitoring of patient responses. AI’s ability to manage and analyze large volumes of clinical trial data in real-time contributed to the swift authorization of vaccines under emergency use authorizations (EUAs). This adaptability and efficiency were critical in meeting the urgent demands of the COVID-19 pandemic while maintaining high standards for safety and efficacy.

Moreover, AI proved essential in tackling logistical challenges during vaccine distribution, particularly in managing the complex cold-chain requirements for mRNA vaccines. AI-powered models optimized vaccine delivery routes, ensuring that doses were efficiently distributed, especially to underserved regions, while minimizing wastage. AI’s role in real-time reallocation of vaccine doses based on population demand and regional infection rates helped to streamline the global rollout, contributing to a more equitable response.

Overall, AI has demonstrated itself to be indispensable in both accelerating vaccine development and ensuring the efficient and timely distribution of vaccines. The successful integration of AI technologies during the COVID-19 pandemic sets a compelling precedent for their continued application in future vaccine research and public health interventions. AI’s capacity to enhance pandemic preparedness, particularly by addressing logistical hurdles and distribution challenges, positions it as a cornerstone of global health strategies going forward.

## Discussion

### Interpretation of Findings

The integration of artificial intelligence (AI) into vaccine development represents a paradigm shift, particularly during the COVID-19 pandemic. This systematic review highlights how AI-driven technologies dramatically accelerated vaccine candidate identification, optimized clinical trial designs, and resolved logistical challenges in manufacturing and distribution. AI’s transformative role in reducing vaccine development timelines—from the standard years to just a few months—was evident in the success of mRNA vaccines, such as Pfizer-BioNTech and Moderna. AI-powered predictive analytics not only enhanced precision in selecting vaccine candidates but also enabled real-time monitoring of clinical trials. This real-time capability optimized patient recruitment, reduced adverse events, and facilitated dynamic trial management, all of which were critical for the rapid approval of COVID-19 vaccines.

Moreover, our review identifies important lessons from the pandemic. Foremost among these is the value of interdisciplinary collaboration between AI researchers, immunologists, and public health experts. AI’s ability to process vast, heterogeneous datasets offers a scalable model for future vaccine development, particularly against emerging infectious diseases. AI’s impact will likely extend beyond vaccines, paving the way for broader applications in therapeutic discovery and precision medicine.

### Challenges and Limitations in AI-Driven Vaccine Development

Despite its remarkable contributions, AI-driven vaccine development faces several challenges and limitations. A primary concern is the quality and diversity of the data used in training AI models. During the COVID-19 pandemic, AI models relied on vast amounts of genetic, clinical, and epidemiological data. However, data availability often varied across regions, leading to potential biases in candidate selection and clinical trial outcomes. This reliance on uneven data poses a risk of skewing AI models, especially when these datasets are not representative of global populations. Additionally, data privacy and security concerns limit AI’s full potential, particularly in healthcare systems where strict regulations govern the sharing of sensitive clinical data.

Another significant limitation is algorithmic bias. AI models trained on incomplete or non-representative datasets risk exacerbating health inequities by favoring certain population groups over others. In the context of vaccine development, this can result in reduced efficacy for underrepresented populations, highlighting the need for more inclusive and diverse datasets. Furthermore, the rapid pace of AI innovation often outpaces regulatory frameworks, leading to challenges in ensuring the ethical use, validation, and public trust in AI-assisted health interventions.

### AI’s Contribution to Future Vaccine Research and Development

AI’s contributions to COVID-19 vaccine development provide a blueprint for future applications in vaccinology. Beyond accelerating vaccine discovery, AI has the potential to revolutionize the design of next-generation vaccines by identifying novel antigens, optimizing adjuvants, and predicting immune responses with unprecedented precision. Additionally, AI could significantly enhance post-marketing surveillance, enabling real-time monitoring of vaccine safety and efficacy across diverse populations. This capability would allow for early detection of adverse events and ensure that vaccine protocols can be swiftly adjusted to maintain public safety.

In future pandemics, AI can offer predictive modeling of disease outbreaks, helping to identify high-risk populations for early vaccination campaigns. By refining machine learning algorithms and expanding the datasets available for training, AI can improve the precision with which vaccines are tailored to specific pathogens and populations. Moreover, AI is poised to contribute to the development of personalized vaccines, which take into account individual genetic profiles and immune responses, thus improving both efficacy and safety.

### Policy Implications and Ethical Considerations

The widespread adoption of AI in vaccine development brings with it significant policy and ethical challenges. Regulatory bodies must adapt to the rapid pace of AI innovation by establishing clear guidelines for the validation and approval of AI-assisted medical interventions. This is especially pertinent in the context of emergency-use authorizations, where expedited approval processes might overlook critical ethical issues, such as informed consent, data privacy, and algorithmic biases.

Additionally, equitable access to AI-driven vaccines is a critical concern. The deployment of AI in resource-rich settings should not marginalize low- and middle-income countries (LMICs), where access to vaccines and advanced AI infrastructure may be limited. Policymakers must establish international frameworks that facilitate the sharing of AI resources, data, and expertise to ensure the equitable distribution of AI-driven innovations. Moreover, ethical oversight is crucial to guarantee that AI models are transparent, accountable, and used responsibly in public health interventions. AI models should be scrutinized to ensure that their predictions and outcomes do not perpetuate or exacerbate existing health inequities.

### Limitations of the Systematic Review

This review has several limitations. The rapidly evolving nature of AI technologies means that some of the latest advancements may not be fully captured in this analysis. Additionally, the heterogeneity of AI applications across different studies made it difficult to conduct a quantitative synthesis, limiting our ability to draw definitive conclusions about AI’s overall effectiveness in vaccine development. Finally, the exclusion of non-English language studies introduces potential selection bias, as significant research may exist in languages other than English that were not included in this review.

## Conclusion

### Summary of Key Lessons from COVID-19 Vaccine Development

The COVID-19 pandemic showcased the transformative capabilities of artificial intelligence (AI) in vaccine development, providing crucial lessons for the future of biomedical research. AI-driven technologies dramatically shortened vaccine development timelines, advancing candidates from conceptualization to clinical deployment in a matter of months—an achievement that typically takes years. Advanced machine learning algorithms enabled the rapid identification of immunogenic epitopes, optimized clinical trial designs, and addressed logistical hurdles in vaccine distribution with unprecedented efficiency.

One of the most profound lessons is AI’s ability to facilitate real-time data analysis and enable adaptive trial designs that adjust dynamically based on interim results. This capacity to process large volumes of clinical and genomic data at high speed will be essential in tackling future pandemics and other global health crises. The pandemic also underscored the critical importance of interdisciplinary collaboration. The seamless integration of AI with immunology, genomics, and public health infrastructure proved key to its success. This collaborative approach should serve as a model for future AI-driven innovations in healthcare and vaccine development.

### Future Directions for AI in Vaccine Research

The future of AI in vaccine research holds extraordinary potential. Beyond reducing development timelines, AI promises to revolutionize the creation of personalized vaccines. By leveraging individual genetic and immunological profiles, AI could enable the design of vaccines tailored to an individual’s immune system, dramatically improving both efficacy and safety.

AI will also play a significant role in post-marketing surveillance, using real-time data to monitor vaccine safety and efficacy across diverse populations. This continuous monitoring can help detect adverse effects earlier and refine vaccine protocols. In the coming years, AI’s reach will likely extend beyond traditional vaccines into fields such as cancer immunotherapy, where personalized vaccines could target specific tumor antigens. Additionally, AI will enhance pandemic preparedness, enabling early modeling of disease outbreaks and optimizing global vaccine distribution strategies.

To fully realize this potential, it is essential that the scientific community invests in developing more robust AI models capable of handling heterogeneous, incomplete datasets— a common challenge in global health. The future of AI in vaccinology lies in refining these models to be more adaptive, inclusive, and capable of accounting for variability across populations.

### Final Thoughts on AI’s Potential in Addressing Global Health Crises

AI represents a paradigm shift in vaccine development, with the potential to radically transform our response to global health emergencies. As demonstrated during the COVID-19 pandemic, AI significantly shortened development timelines, improved the precision of vaccine designs, and optimized large-scale distribution efforts. However, to fully harness AI’s potential, critical challenges remain—most notably ensuring the availability of high-quality data, mitigating algorithmic biases, and establishing ethical frameworks that prioritize transparency and equity in AI-driven healthcare solutions.

In conclusion, AI’s role in vaccine development is just beginning to unfold. The lessons learned from COVID-19 provide a strong foundation for future innovations. By addressing the challenges surrounding AI implementation and fostering continued interdisciplinary collaboration, AI will undoubtedly become an indispensable tool in safeguarding global health and preparing for future pandemics.

**Figure 1:**
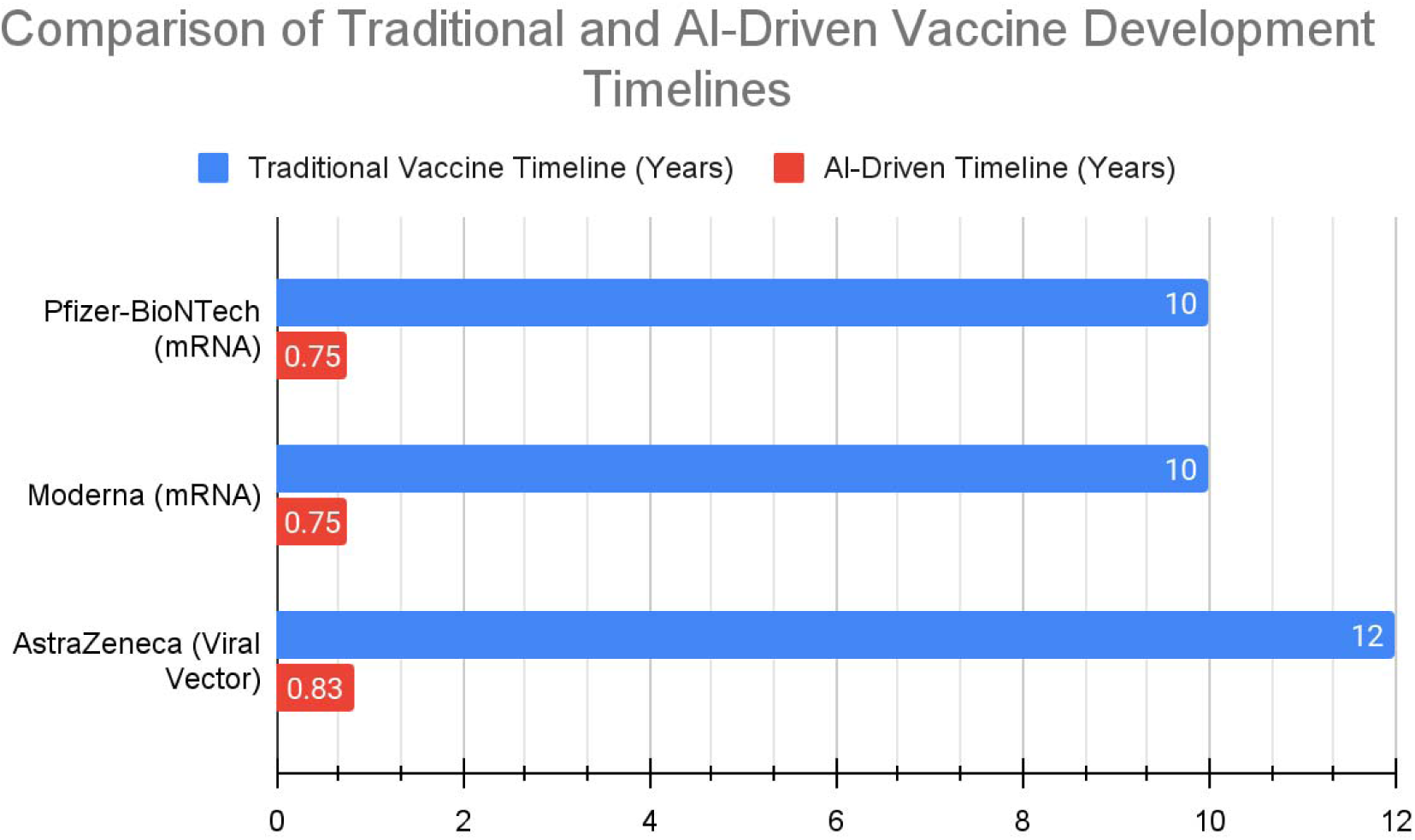
This bar chart illustrates the dramatic reduction in vaccine development timelines achieved through the integration of AI technologies compared to traditional methods. For vaccines such as Pfizer-BioNTech and Moderna, AI-driven approaches reduced timelines from an average of 10 years to just 9 months, showcasing AI’s transformative impact on accelerating both the antigen discovery and clinical trial stages. The chart highlights each key development stage, from antigen discovery to clinical trials, with both AI-accelerated timelines and traditional timelines represented.

**Figure 2:**
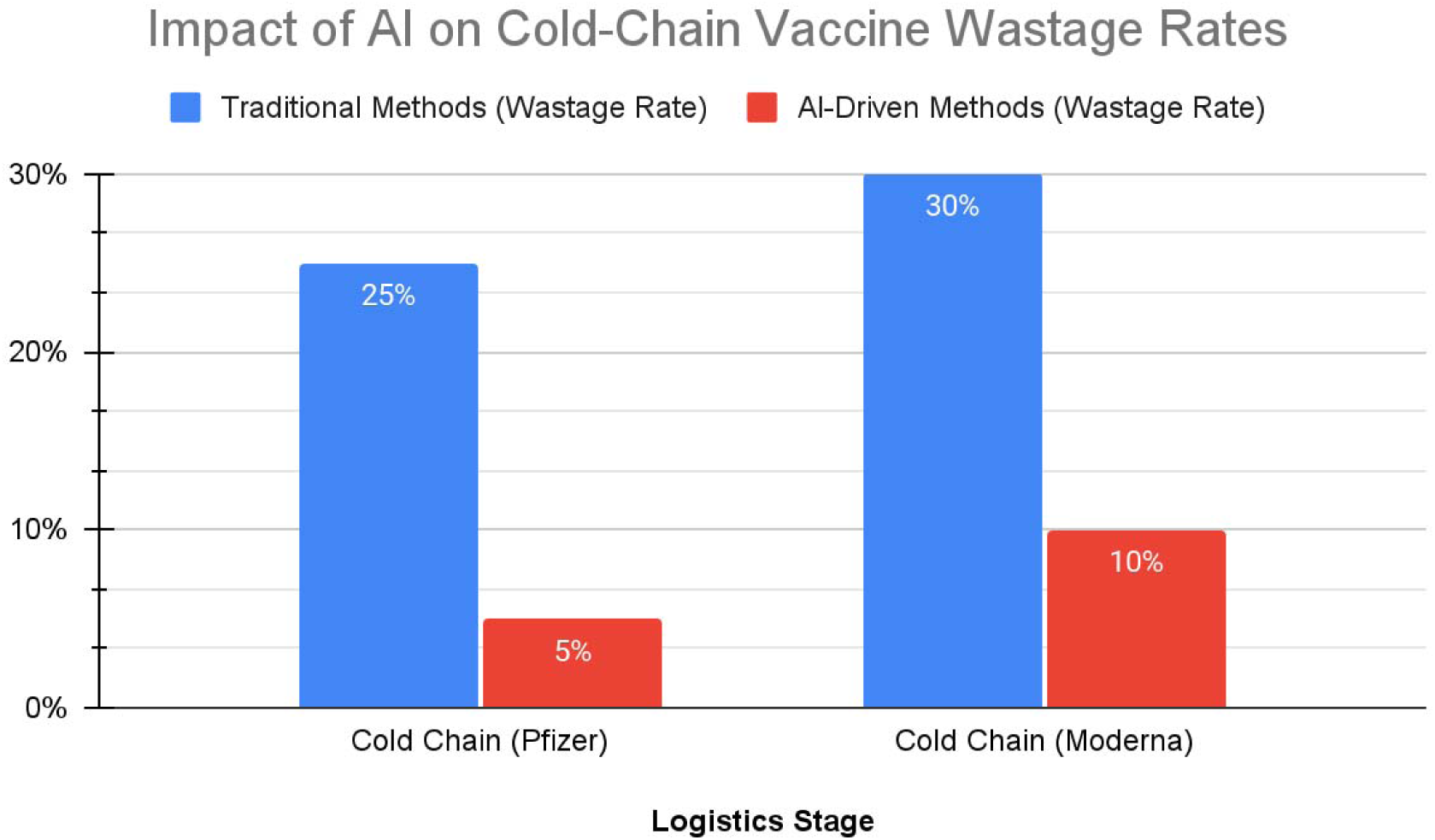
This bar chart compares the vaccine wastage rates due to cold-chain logistics before and after the implementation of AI-driven optimization techniques. In traditional cold-chain management, wastage rates for mRNA vaccines, such as Pfizer and Moderna, were as high as 25-30%, due to inefficiencies in transportation and storage. With the adoption of AI technologies, including real-time monitoring and predictive models, wastage rates dropped significantly to 5-10%, demonstrating AI’s effectiveness in improving logistical processes and reducing vaccine loss during distribution.

**Figure 3:**
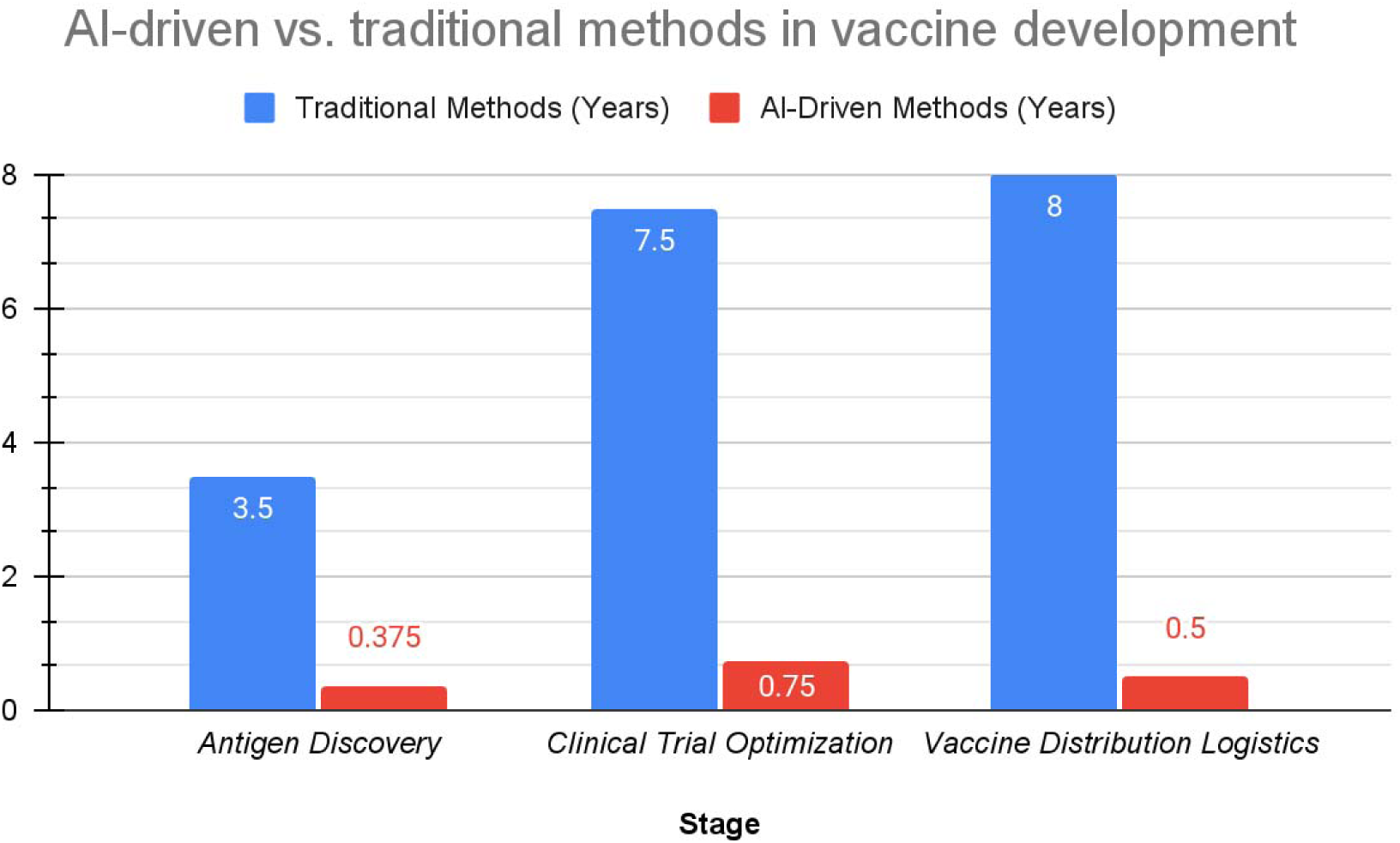
This table summarizes the differences between traditional and AI-driven methods across various stages of vaccine development. It shows how AI models, particularly in antigen discovery and clinical trial design, drastically reduced the time and complexity of these stages compared to traditional methods, cutting years down to months.

## Data Availability

This is a review article and does not report any original research data. All data referenced in this manuscript are derived from previously published studies and publicly available sources, which have been properly cited throughout the text.

## Acknowledgments

The authors, would like to extend sincere gratitude to several individuals and entities who contributed to the completion of this review article. We would like to acknowledge the valuable feedback and guidance provided by our colleagues, who offered critical insights during the conceptualization and development of this manuscript. Special thanks go to the research and technical support teams at the University Lab for their assistance in compiling data and conducting analysis. We also express our deepest appreciation to our parents for their unwavering encouragement and emotional support throughout this research journey, which made this work possible.

## Ethics approval and consent to participate

Not applicable, as this is a review article and does not involve any clinical data, patient participation, or human subjects.

## Consent for publication

Not applicable.

## Availability of Data and Materials

All data generated or analyzed during this study are included in this published article. No new clinical data or patient-related information was generated, as this is a review of existing literature and studies.

## References

1. Thakur, A. (2024). Re-visiting mpox: Stealth Assault on the Brain and Emerging Biomedical Research Insights. Brain Disorders. Available at: https://www.sciencedirect.com/science/article/pii/S2666459324000544

2. Ralbovsky, N. M., Zhang, Y., Williams, D. M. (2024). Machine Learning and Hyperspectral Imaging for Analysis of Human Papillomaviruses (HPV) Vaccine Self-Healing Particles. Analytical Chemistry. Available at: https://pubs.acs.org/doi/abs/10.1021/acs.analchem.4c02327

3. Naseem, A., Rasool, F., Haashmi, F. K., Shoaib, M. H. (2024). Determinants of COVID-19 vaccine hesitancy in university students and support staff in Pakistan: A machine learning and statistical analysis. F1000Research. Available at: https://f1000research.com/articles/13-1241

4. Yao, J., Lin, X., Zhang, X., et al. (2024). Predictive biomarkers for immune checkpoint inhibitors therapy in lung cancer: Using AI in vaccine development. Human Vaccines & Immunotherapeutics. Available at: https://www.tandfonline.com/doi/abs/10.1080/21645515.2024.2406063

5. Ayvaci, M. U. S., Jacobi, V. S., Ryu, Y. (2024). Clinically Guided Adaptive Machine Learning Update Strategies for Predicting Severe COVID-19 Outcomes. The American Journal of Medicine. Available at: https://www.amjmed.com/article/S0002-9343(24)00639-9/abstract

6. Abraham, G., & Choudhary, A. R. (2024). Conservative Management of Diabetic Kidney Disease: Including COVID-19 vaccine implications. Google Books. Available at: https://books.google.com/books?id=8zcqEQAAQBAJ

7. Eviatar, T., Ziv, A., Oved, A., Miller-Barmak, A., & Pappo, A. (2024). Longitudinal safety and efficacy of the BNT162b2 mRNA COVID-19 vaccine in children aged 4–11 years. Vaccine. Available at: https://www.sciencedirect.com/science/article/abs/pii/S0264410X24011083

8. Bollyky, T. (2024). Fauci looks back: Impact of COVID-19 vaccination efforts. The Lancet. Available at: https://www.thelancet.com/journals/lancet/article/PIIS0140-6736(24)02268-2/fulltext

9. Chia, J. E., Faye, M., Jermaine, K., Tesfaye, B., & Modjirom, N. (2024). Navigating the route to polio eradication in the WHO AFRO region: Lessons from COVID-19. The Lancet. Available at: https://www.thelancet.com/journals/lancet/article/PIIS0140-6736(24)02148-2/fulltext

10. Samarasekera, U. (2024). Jean Nachega: Collaborating for research on mpox and COVID-19 in Africa. The Lancet. Available at: https://www.thelancet.com/journals/lancet/article/PIIS0140-6736(24)02267-0/fulltext

11. D. van der Horst, M. E. Carter-Timofte, A. Danneels, et al. (2024). Large-scale Deep Learning Identifies the Antiviral Potential of PKI-179 and MTI-31 Against Coronaviruses. Antiviral Research. Available at: https://www.sciencedirect.com/science/article/pii/S0166354224002213

12. P. Bodapati, E. Zhang, S. Padmanabhan, et al. (2024). A Global Network Analysis of COVID-19 Vaccine Distribution to Predict Breakthrough Cases among the Vaccinated Population. COVID. Available at: https://www.mdpi.com/2673-8112/4/10/107

13. R. Garcia-Carretero, M. Ordoñez-Garcia et al. (2024). Impact and Effectiveness of COVID-19 Vaccines Based on Machine Learning Analysis of a Time Series: A Population-Based Study. Journal of Clinical Medicine. Available at: https://www.mdpi.com/2077-0383/13/19/5890

14. A. Jerfy, O. Selden, et al. (2024). The Growing Impact of Natural Language Processing in Healthcare and Public Health. Inquiry. Available at: https://journals.sagepub.com/doi/abs/10.1177/00469580241290095

15. I. Campo Sánchez-Hermosilla (2024). A Study on NLP Model Ensembles and Data Augmentation Techniques for Separating Critical Thinking from Conspiracy Theories in English Texts. UPM. Available at: https://oa.upm.es/id/eprint/83761

16. S. Chauhan, M. Paliwal, N. Manchanda, et al. (2024). AI and Machine Learning. Google Books. Available at: https://books.google.com/books?hl=en&lr=&id=1J8nEQAAQBAJ&oi=fnd&pg=PA1

17. R. Brent, T. G. McKelvey Jr., J. Matheny (2024). The New Bioweapons: How Synthetic Biology Could Destabilize the World. Foreign Affairs. Available at: https://heinonline.org/hol-cgi-bin/get_pdf.cgi?handle=hein.journals/fora103&section=90

18. P. S. Kumbhar, S. A. Salunkhe (2024). Transformative Impact of Artificial Intelligence on Healthcare. Journal of Computer Based Parallel Processing. Available at: https://matjournals.net/engineering/index.php/JoCPP/article/view/955

19. P. J. Dorsey, C. L. Lau, T. Chang, P. C. Doerschuk et al. (2024). Review of Machine Learning for Lipid Nanoparticle Formulation and Process Development. Journal of Controlled Release. Available at: https://www.sciencedirect.com/science/article/pii/S0022354924004222

20. A. Naseem, F. Rasool, F. K. Haashmi, et al. (2024). Determinants of COVID-19 vaccine hesitancy in university students and support staff in Pakistan. F1000Research. Available at: https://f1000research.com/articles/13-1241

21. L. Peake, G. Adeniyi-Ogunyankin, et al. (2024). Introduction: Gender and Cities. Handbook on Gender and the Urban Space. Available at: https://www.elgaronline.com/edcollchap/book/9781786436139/book-part-9781786436139-6.xml

22. M. A. S. M. Sofian, N. M. Sabri, U. F. M. Bahrin, et al. (2024). Sentiment Analysis on Acceptance of COVID-19 Vaccine for Children based on Support Vector Machine. Applied Sciences and Engineering Technology. Available at: http://semarakilmu.com.my/journals/index.php/applied_sciences_eng_tech/article/view/5449

23. M. V. F. Ferraz, W. C. S. Adan, T. E. Lima, et al. (2024). Design of nanobody targeting SARS-CoV-2 spike glycoprotein using CDR-grafting assisted by molecular simulation and machine learning. bioRxiv. Available at: https://www.biorxiv.org/content/10.1101/2024.09.30.615772.abstract

24. S. N. H. Bukhari, K. A. Ogudo (2024). Prediction of antigenic peptides of SARS-CoV-2 pathogen using machine learning. PeerJ Computer Science. Available at: https://peerj.com/articles/cs-2319/

25. J. Yoo (2024). Cryogenic Electron Microscopy of Infectious Diseases. Frontiers in Molecular Biosciences. Available at: https://www.frontiersin.org/journals/molecular-biosciences/articles/10.3389/fmolb.2024.1506197/full

26. M. M. Ibrahim, E. C. Madu, K. A. Saka et al. (2024). DATA-DRIVEN PARADIGM SHIFTS: Emerging Information Scientists’ Approaches to Information Analysis. Berkeley Journal of Systematic and Management Studies. Available at: https://berkeleypublications.com/bjsmsr/article/view/277

27. J. Tutt, S. Voloshynovskiy (2024). Provable Performance Guarantees of Copy Detection Patterns. arXiv. Available at: https://arxiv.org/abs/2409.17649

28. M. U. S. Ayvaci, V. S. Jacobi, Y. Ryu et al. (2024). Clinically Guided Adaptive Machine Learning Update Strategies for Predicting Severe COVID-19 Outcomes. The American Journal of Medicine. Available at: https://www.amjmed.com/article/S0002-9343(24)00639-9/abstract

29. D. Ravenda, M. M. Valencia-Silva et al. (2024). Social media in healthcare emergency management: insights from Spanish hospitals during the COVID-19 pandemic. International Journal of Emergency Services. Available at: https://www.emerald.com/insight/content/doi/10.1108/IJES-02-2024-0013/full/html

30. S. B. Guo, Y. Meng, L. Lin, et al. (2024). Artificial intelligence alphafold model for molecular biology and drug discovery: a machine-learning-driven informatics investigation. Molecular Cancer. Available at: https://link.springer.com/article/10.1186/s12943-024-02140-6

